# Modeling the impact of vaccination strategies for nursing homes in the context of increased SARS-CoV-2 community transmission and variants

**DOI:** 10.1101/2021.10.25.21265493

**Authors:** Inga Holmdahl, Rebecca Kahn, Kara Jacobs Slifka, Kathleen Dooling, Rachel B. Slayton

**Affiliations:** Center for Communicable Disease Dynamics, Department of Epidemiology, Harvard T.H. Chan School of Public Health, Boston, Massachusetts; COVID-19 Response, US Centers for Disease Control and Prevention, Atlanta, Georgia

## Abstract

Nursing homes (NH) were among the first settings to receive COVID-19 vaccines in the United States, but staff vaccination coverage remains low at an average of 64%. Using an agent-based model, we examined the impact of community prevalence, the Delta variant, staff vaccination coverage, and boosters for residents on outbreak dynamics in nursing homes. We found that increased staff primary series coverage and high booster vaccine effectiveness (VE) in residents leads to fewer infections and that the cumulative incidence is highly dependent on community transmission. Despite high VE, high community transmission resulted in continued symptomatic infections in NHs.

## Background

Nursing home (NH) residents and staff were among the first to receive COVID-19 vaccines, following recommendations from the Centers for Disease Control and Prevention (CDC)’s Advisory Committee on Immunization Practices (ACIP), due to both the high risk of severe disease and the high risk of spread in this congregate setting [1]. Vaccination has contributed to dramatic decreases in overall NH cases; however, in July 2021 cases began increasing again [2]. Recent studies have found decreased vaccine effectiveness (VE) in NHs over time [3], which may be due to waning, decreased effectiveness against the Delta variant, increased importation from the community as COVID-19 incidence has increased, or a combination of these factors.

On September 5, 2021, average vaccination coverage among NH residents was 84% in the United States; however, coverage among NH staff was approximately 64%, with wide variability across facilities [4]. Previous work has shown that vaccine coverage in staff can play a large part in protecting residents against infection [5]. Vaccine mandates have been put in place for many federal government employees and are now being considered for healthcare facilities receiving Centers for Medicare & Medicaid Services (CMS) reimbursement, including NHs [6]. As efforts to increase staff vaccination coverage continue and booster doses for residents are considered, it is important to understand the potential impact of these additional vaccine doses in order to set vaccine program priorities.

Here we examine the impact of high community prevalence and the more infectious Delta variant of SARS-CoV-2 on the expected distribution of infections among vaccinated and unvaccinated people in the NH. We then look at the effect of different vaccination strategies for NH residents and staff, evaluating a range of levels of staff coverage and different scenarios of booster effectiveness.

## Methods

We build upon a previously published model of a NH with 100 residents and 100 staff [5,7]. We incorporate data on resident turnover, with a median stay of 27 days, [8] and keep the NH at 100% capacity. Transmission of SARS-CoV-2 within the nursing home is stochastic, based on the probability of transmission given contact, the number of contacts per day, and the total number of infected individuals (Table S1). Our model assumes all infections are with the Delta variant of SARS-CoV-2, the dominant variant across the US as of August 28, 2021 [9].

Staff have a daily probability of infection from the community, which we vary to reflect different levels of community prevalence. Mandatory non-outbreak screening testing of unvaccinated staff is conducted twice per week for nursing homes located in counties with substantial to high community transmission [10]. When a case is identified, all residents and staff in the nursing home are tested twice weekly for the duration of the outbreak. Reflecting the relaxation of visitation restrictions in nursing homes since March 2021, residents have a daily probability of interacting with a visitor from outside of the nursing home population [11]; each visitor only interacts with a single resident.

We include three vaccine effects of a two-dose vaccine in the model, basing estimates of VE on data from mRNA vaccine studies (Table S1). First, the vaccine confers imperfect protection against infection, reducing the probability of infection given exposure. Second, we incorporate protection against infectiousness (i.e., transmission); this is a measure of the relative infectiousness of vaccinated infected individuals (i.e. breakthrough infections) compared to unvaccinated infected individuals. Third, we model VE against progression to symptoms among vaccinated individuals who get infected (Table S1). VE against symptomatic disease, the primary endpoint in the vaccine trials, is a combination of VE against infection and VE against progression to symptoms. To account for the varied estimates of these key measures from different studies, we vary each of these VE parameters (Table S1). Given the potential impact of age on VE, we compare a scenario in which the VE against infection is lower for residents than for staff [12-14] to a scenario in which it is equal.

We compare two primary vaccination strategies: 1) varying staff vaccination coverage between 40% and 100%, and 2) giving a third dose as a booster to all vaccinated residents (assuming coverage in residents remains constant, at 80%). The effect of boosters on VE against infection is unknown, so we consider a range from 60%-90%. At the beginning of each simulation, we assumed everyone who is vaccinated has already received two doses; in scenarios with a third-dose booster, the booster is given on the first day, and takes 14 days to take effect.

We show results from 100 simulations after two months. Our primary outcomes of interest are the cumulative incidence of symptomatic infections and the cumulative incidence of all infections (symptomatic and asymptomatic) in residents. We also look at the cumulative incidence of symptomatic infections and all infections in the entire nursing home.

## Results

We estimated the distribution of cumulative incidence among residents after a two-month simulation period, disaggregated by symptom status and vaccination status (Figure S1). We focused initially on examining the impact of two key parameters—VE against infection among residents and staff importation rate—in the absence of booster doses. In these simulations, resident vaccination coverage is 80% and staff vaccination coverage is 60%, and there is an average of 217 unique residents in the nursing home over the two-month period. Across the 100 simulations, stochasticity leads to varying numbers of infections; however, we demonstrate important trends by comparing the distribution of infections across simulations. A higher daily importation rate (i.e., more frequent staff infections from the community) results in more infections among both unvaccinated and vaccinated residents. Higher VE against infection among residents leads to similar trends in the distribution of infections by vaccine and symptom status but lower cumulative incidence. While only 20% of residents are unvaccinated, they contribute almost as many symptomatic infections as the 80% of residents who are vaccinated. We found that the majority of infections in vaccinated residents would be asymptomatic due to the vaccine’s efficacy against symptomatic disease.

We evaluated the impact of varying levels of staff coverage and of booster dose effectiveness on the two-month cumulative incidence among residents. In these simulations, we assumed booster doses are provided to all vaccinated residents, including both residents in the nursing home at the beginning of the simulations and incoming residents. We found that increased staff coverage (top of figures) led to fewer infections, and the cumulative incidence was highly dependent on community transmission (left vs. right panels). Providing boosters to residents led to fewer infections, with the magnitude of the impact of boosters increasing with higher VE (right of figures). The trends were similar when comparing the cumulative incidence of symptomatic infections (Figure 1a) with cumulative incidence of all infections (Figure 1b).

**Figure 1.**
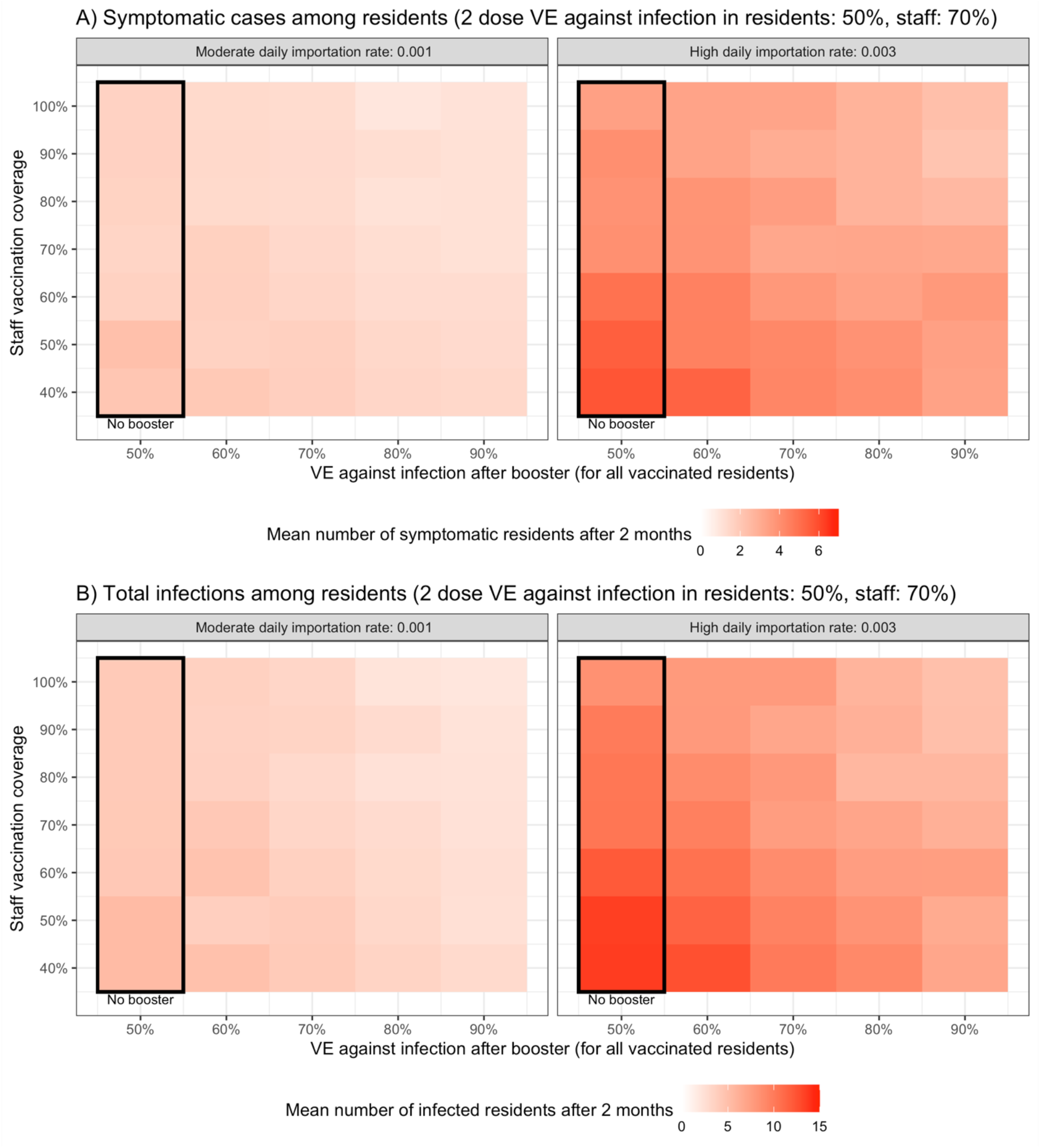
Average cumulative number of infections across 100 simulations of A) symptomatic residents and B) residents (symptomatic and asymptomatic) after 2 months, varying staff coverage (rows), booster VE (columns), and staff importation rates (panels)

We observed similar trends when looking at symptomatic infections (Figure S2a) and total infections (Figure S2b) in residents and staff as compared to in residents only. However, in these simulations, there was a larger impact of increasing staff coverage as these metrics capture the direct protection of the vaccine in staff in addition to the indirect protection provided to residents. When VE against infection in residents was equivalent to VE in staff (Figure S3, S4), we saw similar trends but lower cumulative incidence. When we assume that VE against infectiousness is higher—i.e., infected vaccinated individuals have even lower infectiousness than in the baseline simulations—we also saw similar trends but lower cumulative incidence (Figure S5, S6).

## Discussion

Several factors influence the risk of outbreaks of SARS-CoV-2 in nursing homes, and multifaceted approaches are required to protect this vulnerable population at high risk of severe outcomes. We find that maximizing vaccine coverage among nursing home staff remains a critical tool for preventing infections, supporting pending CMS mandates [15]. While boosters for residents can help reduce infections, our simulations show the magnitude of their effect depends on their efficacy. Early evidence from Israel suggests boosters increase VE in people aged 60 and older, although the durability of this increased protection, particularly in this vulnerable population, is unknown [16,17]. However, even with high efficacy boosters, when community transmission is high, our simulations suggest that symptomatic infections in nursing homes will continue.

Community transmission is one of the main drivers of SARS-CoV-2 infections in nursing homes. The epidemic curves in nursing homes and in the United States follow similar patterns, highlighting that control of community transmission and continued infection prevention and control measures are important for protecting the nursing home population. Our results demonstrate that an increase in nursing home infections does not necessarily indicate lower or waning VE if community transmission is rising.

The largest reduction in transmission occurred in simulations with both high staff coverage and high booster efficacy, although each scenario on its own also led to reduced transmission. Direct comparison of the two strategies is challenging because the impact of boosters depends on booster efficacy. Additionally, the two strategies require different numbers of vaccines; increasing staff COVID-19 vaccination coverage will require fewer doses in most nursing homes than providing boosters to all vaccinated residents, particularly given the high rate of resident turnover.

Our results are subject to several limitations. As described previously [5,7], while our model incorporates data on contact structure from nursing homes and separately models contacts between residents and staff, we do not further differentiate between types of contacts. We also do not incorporate any prior infection; we assume that all residents and staff have never been infected, which may underestimate the level of immune protection (particularly for those who are unvaccinated) in a setting with high rates of past infection. Although they are important endpoints, given limited data and heterogeneity in hospitalization criteria between facilities, we do not model hospitalizations and deaths and instead only distinguish between asymptomatic and symptomatic infection. This may underestimate the impact of vaccines on disease severity, as symptomatic infections among vaccinated individuals may be less severe. Finally, we do not explore the impact of supply shortages in this model. Given the high rate of resident turnover, increasing staff COVID-19 vaccination coverage will require fewer doses in most nursing homes than providing boosters to all vaccinated residents. This may be an important consideration, in situations with limited vaccine supply.

## Data Availability

Code will be available on github.

https://github.com/rek160/NHboosters

## Disclaimer

The findings and conclusions in this report are those of the authors and do not necessarily represent the official position of the Centers for Disease Control and Prevention.

## Conflicts of Interest

RK discloses consulting fees from the Pan American Health Organization.

## Funding

RK was supported by the National Cancer Institute U01: U01 CA261277.

## Supplement

**Table S1.**

| Parameter | Values** |
| --- | --- |
| $R_0$ | 6 (Delta estimate) [1,2] |
| Probability of infection per infectious contact | 0.0372 |
| Daily contacts staff-staff | 2 [3] |
| Daily contacts residents - staff | 6 [3,4] |
| Daily contacts staff - residents | 6 - 12 [3] |
| Daily contacts residents - residents (non-roommates) | 6 |
| Daily visitor probability | 0.1 |
| Latent period (days) | 3-5 [5] |
| Proportion of unvaccinated staff asymptomatic | 0.4 [6,7] |
| Proportion of unvaccinated residents asymptomatic | 0.2 [8-10] |
| Duration of pre-symptomatic transmission (days) | 2 [5,7,11] |
| Time in infectious compartment (days); infectiousness dependent on viral load | 14 [12] |
| Reduction in force of infection per contact from PPE | 95% [13] |
| COVID-19 mortality (daily) | 0.02 |
| Mean peak viral load (copies/mL) | $10^8$ [14] |
| Limit of detection - rapid antigen test (copies/mL) | $10^5$ [15-18] |
| Antigen test specificity | 1 |
| Viral load threshold for infectiousness (copies/mL) | $10^4$ |
| Viral load threshold for high infectiousness (copies/mL) | $10^7$ |
| Time until effect of vaccine dose after vaccination (days) | 14 days |
| Staff daily probability of community infection | 0.001 - 0.003 [19] |
| Resident vaccination coverage | 80% [20] |
| Staff vaccination coverage | 40% - 100% |
| 2 dose vaccine efficacy against infection (staff) | 70% [21, 22] |
| 2 dose vaccine efficacy against infection (residents) | 50% - 70% [21, 22] |
| 2 dose vaccine efficacy against progression to symptoms (staff)* | 70% [22] |
| 2 dose vaccine efficacy against progression to symptoms (resident)* | 60% - 66.67% [22] |
| 2 dose vaccine efficacy against symptomatic disease* (staff) | 90% [22] |
| 2 dose vaccine efficacy against symptomatic disease* (residents) | 80% - 90% [21, 22] |
| vaccine efficacy against infectiousness (residents and staff) | 50% - 90% |
| Booster dose vaccine efficacy against infection (residents, takes 2 weeks to take effect) | 60% - 90% |

\* VE against symptomatic disease = $1 - (1 - \text{VE against infection}) * (1 - \text{VE against progression to symptoms})$
\*\* If no references are listed, values are assumed

**Figure S1.**
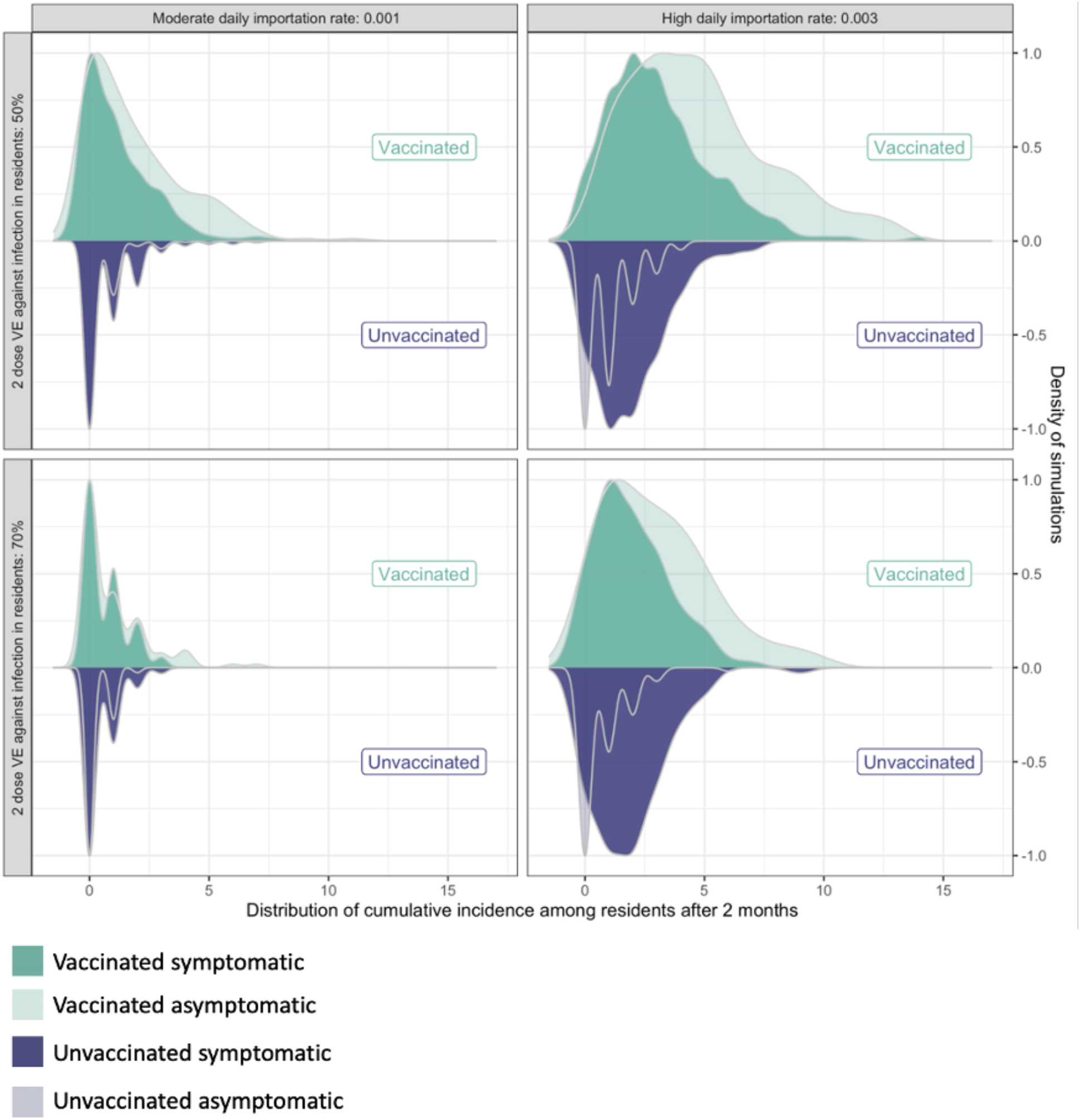
Distribution of cumulative cases across 100 simulations over a two-month period disaggregated by symptom and vaccination status. Over this period there are an average of 217 unique residents in the nursing home; 80% of residents are fully vaccinated (including those who are admitted during the simulation) and 60% of staff are fully vaccinated.

**Figure S2.**
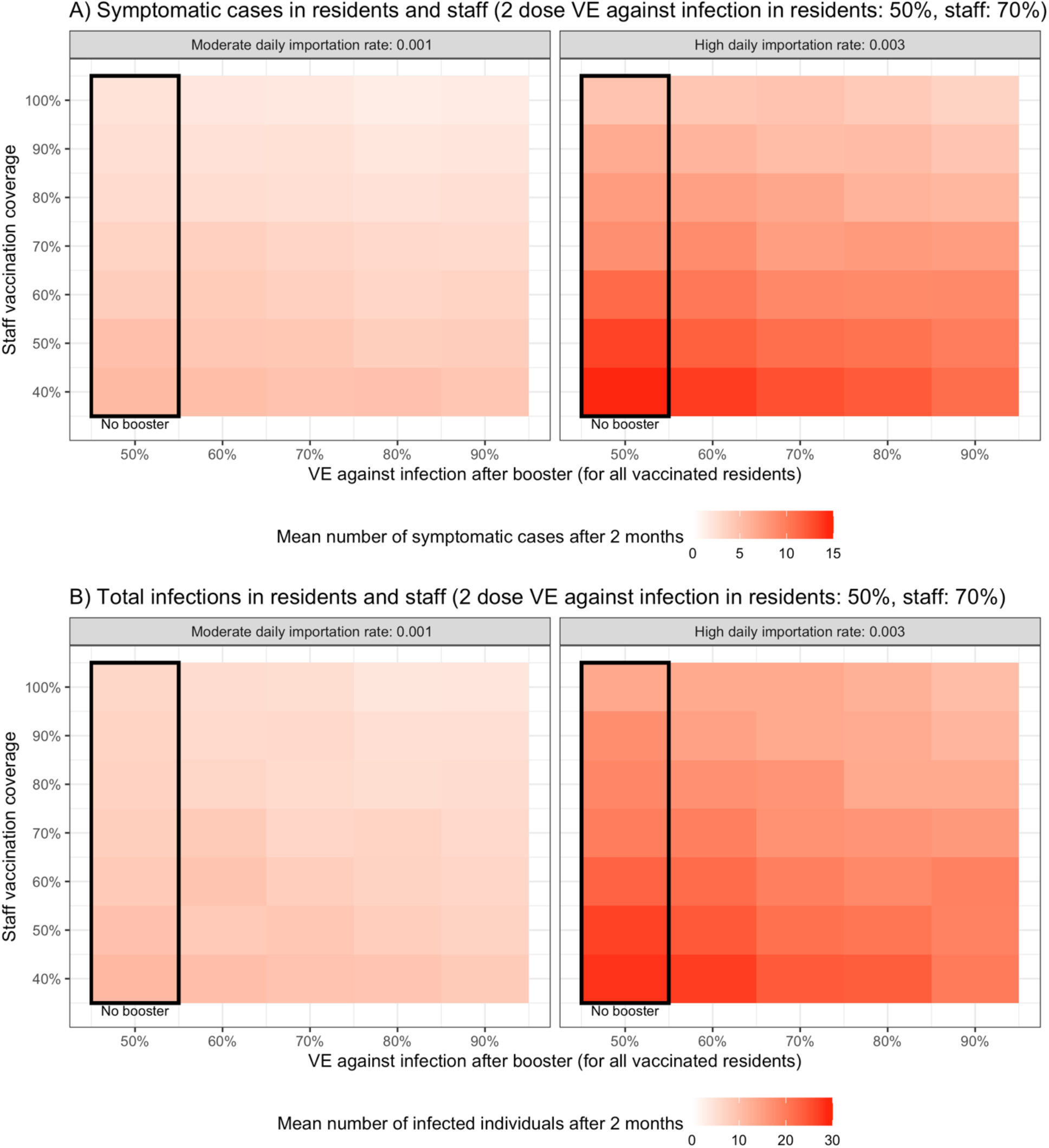
Average cumulative number across 100 simulations of A) symptomatic residents and staff and B) infected residents and staff (symptomatic and asymptomatic) after 2 months, varying staff coverage (rows), booster VE (columns), and staff importation rates (panels)

**Figure S3.**
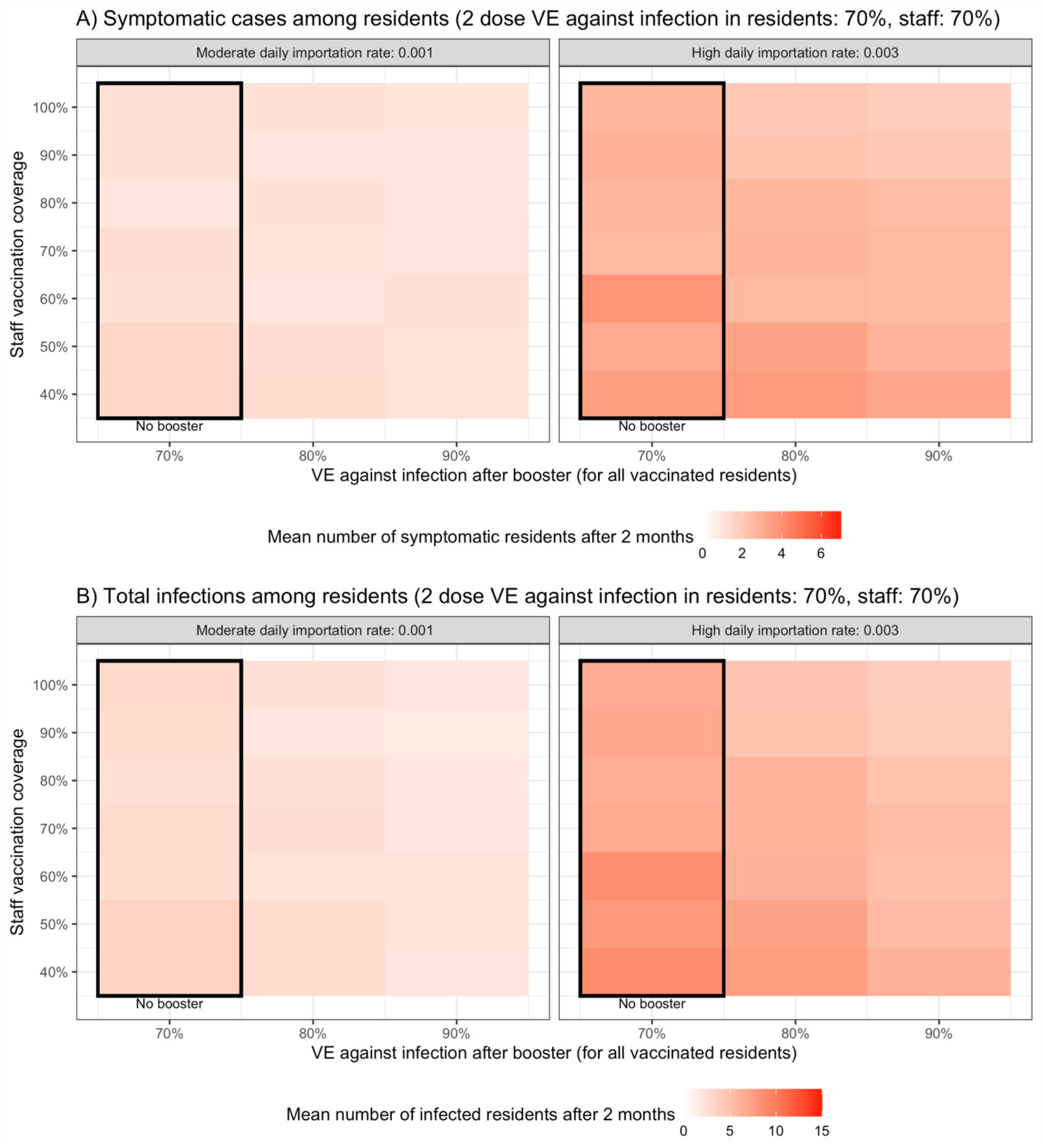
Average cumulative number across 100 simulations of A) symptomatic residents and B) infected residents (symptomatic and asymptomatic) after 2 months, varying staff coverage (rows), booster VE (columns), and staff importation rates (panels) in simulations with higher VE against infection among residents than in the baseline simulations

**Figure S4.**
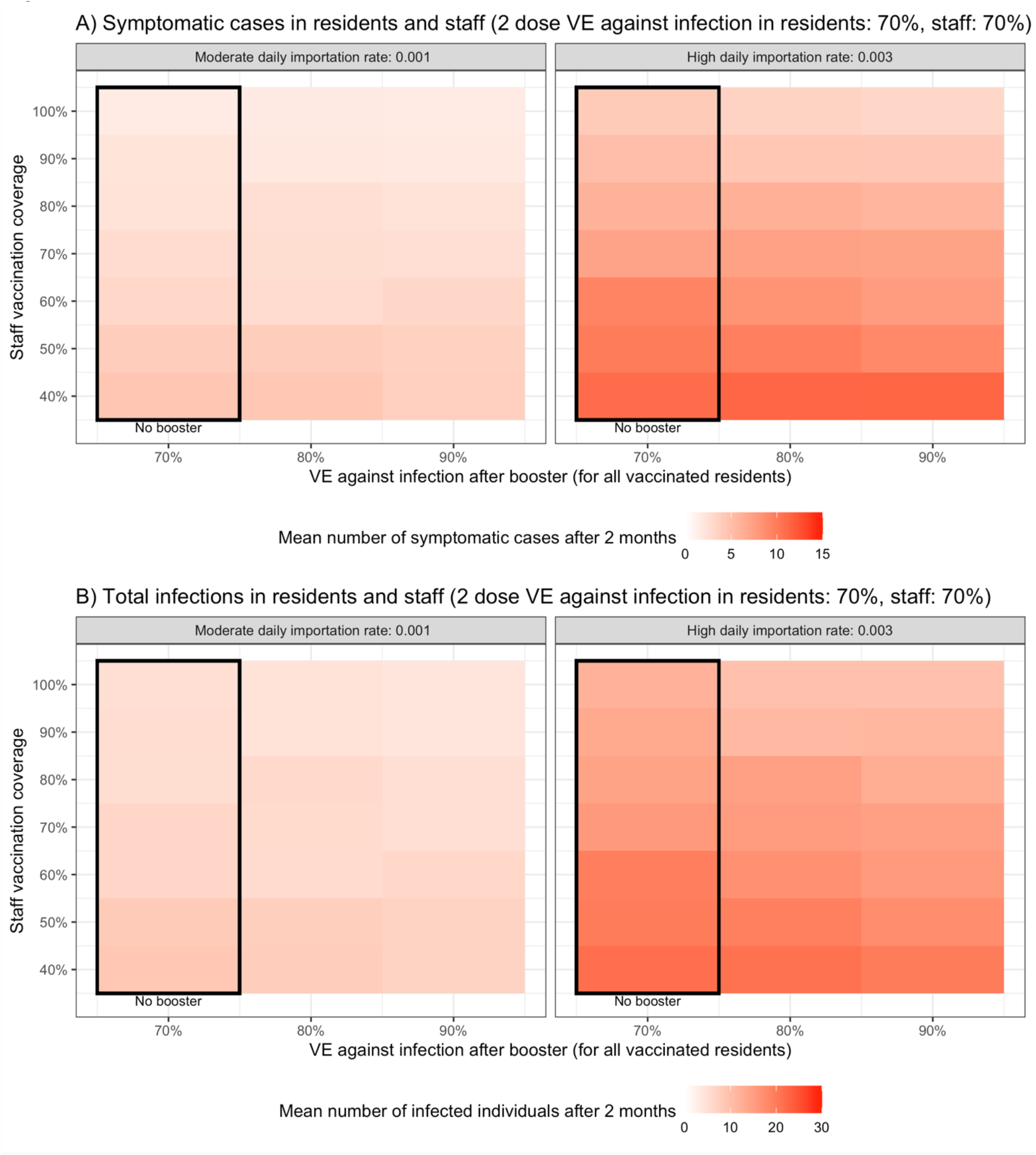
Average cumulative number across 100 simulations of A) symptomatic residents and staff and B) infected residents and staff (symptomatic and asymptomatic) after 2 months, varying staff coverage (rows), booster VE (columns), and staff importation rates (panels) in simulations with higher VE against infection among residents than in the baseline simulations

**Figure S5.**
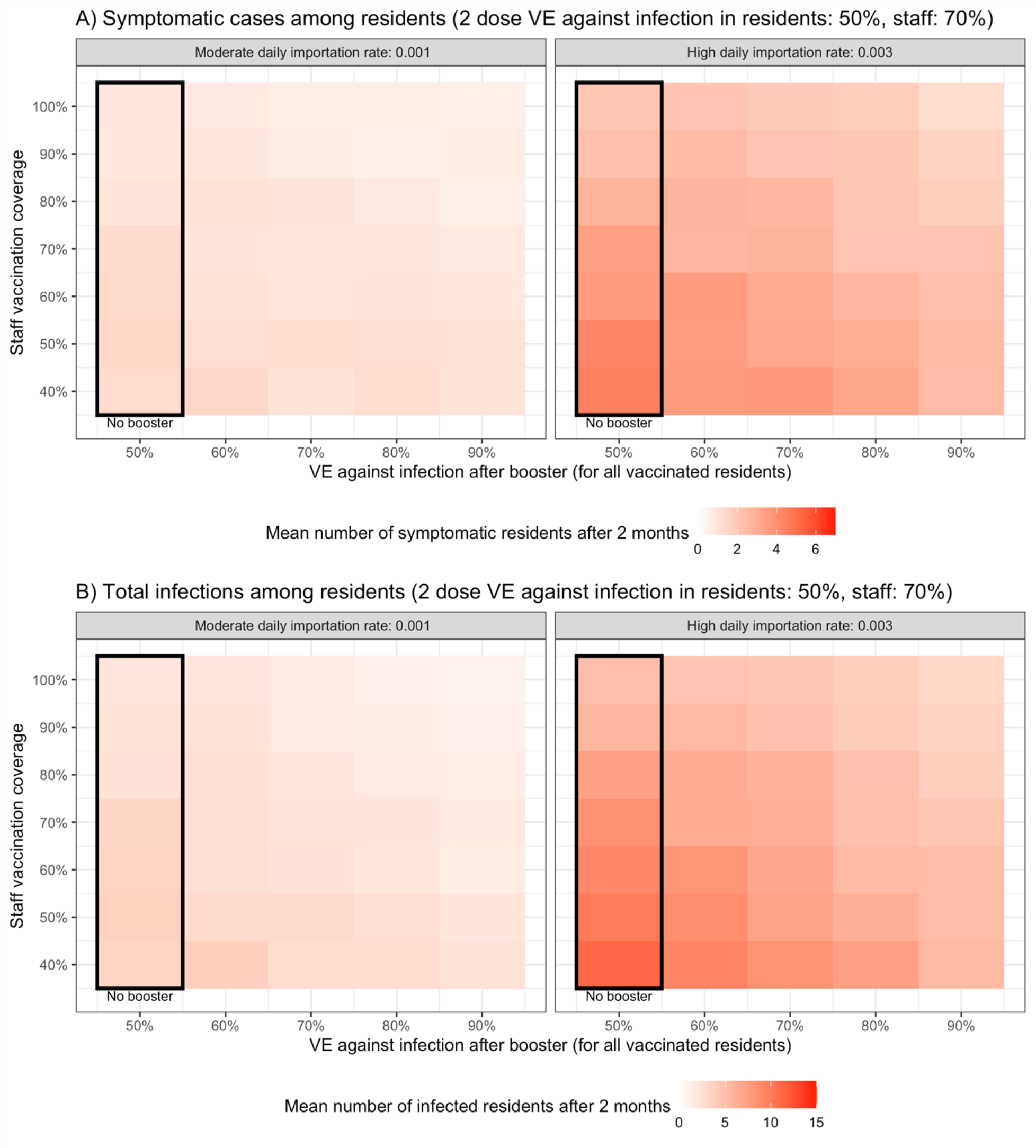
Average cumulative number across 100 simulations of A) symptomatic residents and B) infected residents (symptomatic and asymptomatic) after 2 months, varying staff coverage (rows), booster VE (columns), and staff importation rates (panels) in simulations with higher VE against infectiousness among residents than in the baseline simulations (90% vs. 50%)

**Figure S6.**
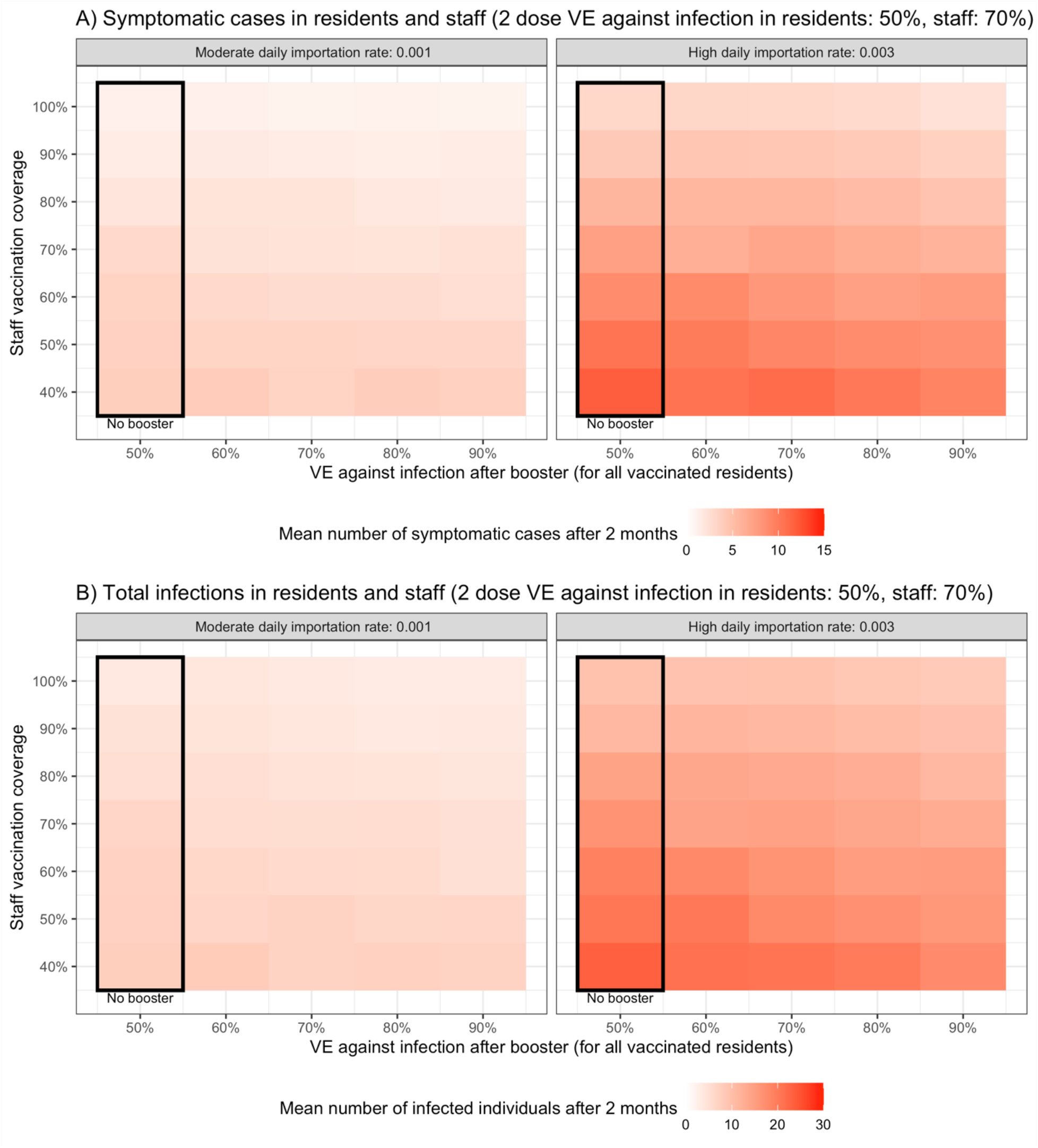
Average cumulative number across 100 simulations of A) symptomatic residents and staff and B) infected residents and staff (symptomatic and asymptomatic) after 2 months, varying staff coverage (rows), booster VE (columns), and staff importation rates (panels) in simulations with higher VE against infectiousness among residents than in the baseline simulations (90% vs. 50%)

## References

1. Dooling K. The Advisory Committee on Immunization Practices’ Updated Interim Recommendation for Allocation of COVID-19 Vaccine — United States, December 2020. MMWR Morb Mortal Wkly Rep 2021; 69. Available at: https://www.cdc.gov/mmwr/volumes/69/wr/mm695152e2.htm. Accessed 5 September 2021.

2. CDC. National Healthcare Safety Network COVID-19 Data Dashboard. 2021. Available at: https://www.cdc.gov/nhsn/covid19/ltc-report-overview.html. Accessed 5 September 2021.

3. Nanduri S. Effectiveness of Pfizer-BioNTech and Moderna Vaccines in Preventing SARS-CoV-2 Infection Among Nursing Home Residents Before and During Widespread Circulation of the SARS-CoV-2 B.1.617.2 (Delta) Variant — National Healthcare Safety Network, March 1–August 1, 2021. MMWR Morb Mortal Wkly Rep 2021; 70. Available at: https://www.cdc.gov/mmwr/volumes/70/wr/mm7034e3.htm. Accessed 5 September 2021.

4. Centers for Medicare & Medicaid Services Data. Available at: https://www.google.com/url?q=https://data.cms.gov/covid-19/covid-19-nursing-home-data&sa=D&source=editors&ust=1630867449026000&usg=AOvVaw3FTiBsOErdqOXvluNCr4Cz. Accessed 5 September 2021.

5. Kahn R, Holmdahl I, Reddy S, Jernigan J, Mina MJ, Slayton RB. Mathematical Modeling to Inform Vaccination Strategies and Testing Approaches for Coronavirus Disease 2019 (COVID-19) in Nursing Homes. Clinical Infectious Diseases. 2021; Available at: http://dx.doi.org/10.1093/cid/ciab517.

6. President Biden’s COVID-19 Plan. 2021. Available at: https://www.whitehouse.gov/covidplan/. Accessed 15 September 2021.

7. Holmdahl I, Kahn R, Hay JA, Buckee CO, Mina MJ. Estimation of Transmission of COVID-19 in Simulated Nursing Homes With Frequent Testing and Immunity-Based Staffing. JAMA Netw Open 2021; 4:e2110071–e2110071.

8. Minimum Data Set 3.0 Public Reports. Available at: https://www.cms.gov/Research-Statistics-Data-and-Systems/Computer-Data-and-Systems/Minimum-Data-Set-3-0-Public-Reports. Accessed 5 September 2021.

9. CDC. COVID Data Tracker. 2020. Available at: https://covid.cdc.gov/covid-data-tracker/. Accessed 5 September 2021.

10. Memorandum QSO-20-38-NH. CMS, 2021 Available at: https://www.cms.gov/files/document/qso-20-38-nh-revised.pdf.

11. Memorandum QSO-20-39-NH. CMS, 2021 Available at: https://www.cms.gov/files/document/qso-20-39-nh-revised.pdf.

12. Collier DA, Ferreira iatm, Kotagiri P, et al. Age-related immune response heterogeneity to SARS-CoV-2 vaccine BNT162b2. Nature 2021; 596:417–422.

13. Pouwels KB, Pritchard E, Matthews PC, et al. Impact of Delta on viral burden and vaccine effectiveness against new SARS-CoV-2 infections in the UK. medRxiv 2021; :2021.08.18.21262237.

14. Abe KT, Hu Q, Mozafarihashjin M, et al. Neutralizing antibody responses to SARS-CoV-2 variants in vaccinated Ontario long-term care home residents and workers. medRxiv 2021; :2021.08.06.21261721.

15. Biden-Harris Administration Takes Additional Action to Protect America’s Nursing Home Residents from COVID-19. Available at: https://www.cms.gov/newsroom/press-releases/biden-harris-administration-takes-additional-action-protect-americas-nursing-home-residents-covid-19. Accessed 9 September 2021.

16. Bar-On YM, Goldberg Y, Mandel M, et al. BNT162b2 vaccine booster dose protection: A nationwide study from Israel. medRxiv 2021; :2021.08.27.21262679.

17. Patalon T, Gazit S, Pitzer VE, Prunas O, Warren JL, Weinberger DM. Short Term Reduction in the Odds of Testing Positive for SARS-CoV-2; a Comparison Between Two Doses and Three doses of the BNT162b2 Vaccine. medRxiv 2021; :2021.08.29.21262792.

## References

1. CDC. COVID Data Tracker. 2020. Available at: https://covid.cdc.gov/covid-data-tracker/. Accessed 5 September 2021.

2. Burki TK. Lifting of COVID-19 restrictions in the UK and the Delta variant. Lancet Respir Med 2021; 9:e85.

3. Anglo R. Chief Clinical Officer, Chelsea Jewish Life Care, Personal communication, Jun 10, 2020.

4. Samore M. Professor of Internal Medicine, University of Utah. Personal communication, Dec 08, 2020.

5. Bar-On YM, Flamholz A, Phillips R, Milo R. SARS-CoV-2 (COVID-19) by the numbers. Elife 2020; 9. Available at: https://www.ncbi.nlm.nih.gov/pmc/articles/PMC7224694/. Accessed 27 September 2020.

6. Oran DP, Topol EJ. Prevalence of Asymptomatic SARS-CoV-2 Infection : A Narrative Review. Ann Intern Med 2020; 173:362–367.

7. Johansson MA, Quandelacy TM, Kada S, et al. SARS-CoV-2 Transmission From People Without COVID-19 Symptoms. JAMA Netw Open 2021; 4:e2035057–e2035057.

8. CDC. Healthcare Workers. 2020. Available at: https://www.cdc.gov/coronavirus/2019-ncov/hcp/planning-scenarios.html. Accessed 17 January 2021.

9. Lennon NJ, Bhattacharyya RP, Mina MJ, et al. Comparison of viral levels in individuals with or without symptoms at time of COVID-19 testing among 32,480 residents and staff of nursing homes and assisted living facilities in Massachusetts. Public and Global Health. 2020; Available at: https://www.medrxiv.org/content/10.1101/2020.07.20.20157792v1.abstract.

10. Byambasuren O, Cardona M, Bell K, Clark J, McLaws M-L, Glasziou P. Estimating the extent of asymptomatic COVID-19 and its potential for community transmission: systematic review and meta-analysis. Available at: http://dx.doi.org/10.1101/2020.05.10.20097543.

11. Li Q, Guan X, Wu P, et al. Early Transmission Dynamics in Wuhan, China, of Novel Coronavirus-Infected Pneumonia. N Engl J Med 2020; 382:1199–1207.

12. Holmdahl I, Kahn R, Hay JA, Buckee CO, Mina MJ. Estimation of Transmission of COVID-19 in Simulated Nursing Homes With Frequent Testing and Immunity-Based Staffing. JAMA Open 2021; 4:e2110071–e2110071.

13. Livingston E, Desai A, Berkwits M. Sourcing Personal Protective Equipment During the COVID-19 Pandemic. JAMA 2020; 323:1912–1914.

14. Wölfel R, Corman VM, Guggemos W, et al. Virological assessment of hospitalized patients with COVID-2019. Nature 2020; 581:465–469.

15. Larremore DB, Wilder B, Lester E, et al. Test sensitivity is secondary to frequency and turnaround time for COVID-19 surveillance. medRxiv 2020; :2020.06.22.20136309.

16. Butler DJ, Mozsary C, Meydan C, et al. Shotgun Transcriptome and Isothermal Profiling of SARS-CoV-2 Infection Reveals Unique Host Responses, Viral Diversification, and Drug Interactions. bioRxiv 2020; Available at: http://dx.doi.org/10.1101/2020.04.20.048066.

17. Dao Thi VL, Herbst K, Boerner K, et al. A colorimetric RT-LAMP assay and LAMP-sequencing for detecting SARS-CoV-2 RNA in clinical samples. Sci Transl Med 2020; 12. Available at: http://dx.doi.org/10.1126/scitranslmed.abc7075.

18. Meyerson NR, Yang Q, Clark SK, et al. A community-deployable SARS-CoV-2 screening test using raw saliva with 45 minutes sample-to-results turnaround. Infectious Diseases (except HIV/AIDS). 2020; Available at: https://www.medrxiv.org/content/10.1101/2020.07.16.20150250v1.abstract.

19. CDC. National Healthcare Safety Network COVID-19 Data Dashboard. 2021. Available at: https://www.cdc.gov/nhsn/covid19/ltc-report-overview.html. Accessed 5 September 2021.

20. Centers for Medicare & Medicaid Services Data. Available at: https://www.google.com/url?q=https://data.cms.gov/covid-19/covid-19-nursing-home-data&sa=D&source=editors&ust=1630867449026000&usg=AOvVaw3FTiBsOErdqOXvluNCr4Cz. Accessed 5 September 2021.

21. Nanduri S. Effectiveness of Pfizer-BioNTech and Moderna Vaccines in Preventing SARS-CoV-2 Infection Among Nursing Home Residents Before and During Widespread Circulation of the SARS-CoV-2 B.1.617.2 (Delta) Variant — National Healthcare Safety Network, March 1–August 1, 2021. MMWR Morb Mortal Wkly Rep 2021; 70. Available at: https://www.cdc.gov/mmwr/volumes/70/wr/mm7034e3.htm. Accessed 5 September 2021.

22. Higdon MM, Wahl B, Jones CB, et al. A systematic review of COVID-19 vaccine efficacy and effectiveness against SARS-CoV-2 infection and disease. medRxiv 2021; :2021.09.17.21263549.

